# Germline predisposition to oncogenic alkylating damage in colorectal cancer

**DOI:** 10.1101/2025.06.30.25330572

**Authors:** Carino Gurjao, Jules Cazaubiel, Chichun Tan, Brendan Reardon, Matan Hofree, Tomotaka Ugai, Jeffrey A. Meyerhardt, Jonathan A. Nowak, Edward L. Giovannucci, Jeffrey P. Townsend, Shuji Ogino, Marios Giannakis

## Abstract

**Background:** Red meat consumption is a risk factor for colorectal cancer (CRC) and has been linked to tumor alkylating DNA damage. *rs16906252*-T is a cis expression quantitative trait locus (eQTL) variant associated with silencing of *MGMT*, a central alkylating damage repair gene. We hypothesize that *rs16906252*-T carriers are predisposed to alkylating damage mutations.

**Methods:** We conducted mutational signature deconvolution of CRC whole-exome sequencing data from The Cancer Genome Atlas (TCGA, n = 540), the Nurses’ Health Studies/ Health Professional Follow-up Study (NHS/HPFS, n = 900) as well as non-western samples from the Pan-Cancer Analysis of Whole Genomes (COCA-CN, n = 295); and examined the relationship of *rs16906252*-T with putative alkylation-dependent tumor mutations. Leveraging lifestyle data from NHS/HPFS, we also investigated the interaction between red meat consumption and *rs16906252*-T.

**Results:** Among CRC patients, *rs16906252*-T carriers exhibited higher tumor alkylating damage compared to non-carriers. In the general population*, rs16906252*-T is largely absent in individuals with East Asian ancestries, and we consistently find a negligible contribution of alkylating damage in CRC patients with East Asian ancestries. We show that the alkylating mutational signature’s carcinogenicity is mainly mediated by *KRAS* G12D and G13D mutations. We also observe a synergistic effect of *rs16906252*-T with high pre-diagnosis red meat intake for tumor alkylating damage.

**Conclusions:** *MGMT rs16906252*-T carriers are predisposed to CRC oncogenic alkylating damage which is potentiated by red meat intake.

**Impact:** Our results support a causal relationship between red meat and CRC and can lead to tailored dietary and screening guidelines for CRC prevention.

## Introduction

Gene variants can have a wide range of effects on cancer initiation and development. High-penetrance variants can cause familial cancers such as those associated with Lynch syndrome or *MUTYH*-associated polyposis^1^, but are rare and affect initiation of only a small percentage of all colorectal cancers (CRC). On the other hand, low-penetrance variants can be maintained at higher frequency and affect a larger segment of the population, contributing to cancer risk by synergistic effects with carcinogenic exposures^2,3^. These gene–environment (G×E) interactions that influence cancer risk have frequently been thought to function by facilitating the acquisition of somatic mutations required for cancer initiation and progression. Such G×E-associated variants can be detected in Genome-Wide Association Studies (GWAS) but have not previously been associated with carcinogenic exposures in CRC and the consequent pathogenic mutations. Recent advances in mutation analysis have made it possible to deconvolute signatures of mutational processes that tumor cells experienced prior to sequencing. These mutational signatures can provide insights into the carcinogenic activity of mutagenic exposures and have made it possible to measure the amount of DNA damage stemming from specific exposures. Linking such mutational signatures to germline variants provides compelling evidence regarding the etiology of exposure and the nature of variation in susceptibility^4^.

We previously discovered a CRC alkylating mutational signature that was associated with high pre-diagnosis red-meat intake^5^. O-6-methylguanine-DNA methyltransferase (MGMT) is a major DNA-damage repair protein that removes alkylating damage, notably methylated guanine residues^6^. The *rs16906252-T (NM_002412.2:c.-56C>T) MGMT* promoter variant is a cis expression quantitative trait locus (eQTL) associated with *MGMT* promoter CpG-island hypermethylation and gene silencing^7,8^. This hypermethylation and gene silencing is observed to occur across diverse normal tissues, including blood samples and healthy colonic tissues from the Genotype-Tissue Expression^9^ database (GTEx, **Figure S1A**). This strong association with gene silencing was also previously observed across cancers—including CRC^6,10^. The *rs16906252*-T *MGMT* promoter variant has also been identified as a risk factor for CRC^11^, supported by a large Genome-Wide Association Study of DNA repair genes^7^ (*n* = 11,114 CRCs and 14,659 controls; **Figure S1B**). Given the strong association of *rs16906252*-T with silencing of the alkylating damage-repair gene *MGMT*, we sought to explore the relationship between this variant and the CRC alkylating mutational signature.

## Materials and Methods

### Ethics Statement

Institutional review board approval and written informed consent were obtained for all patients in the original published studies. All of the tumor data is publicly available via dbGaP (https://dbgap.ncbi.nlm.nih.gov/aa/wga.cgi) and the ICGC Data Access Compliance Office (DACO; http://icgc.org/daco).

### Study populations

We utilized data from three prospective cohort studies in the U.S., the Nurses’ Health Study I, the Nurses’ Health Study II, and the Health Professionals Follow-up Study (HPFS, including 51,529 men aged 40–75 years followed since 1986)^12^. The study participants were sent questionnaires biennially to update information on lifestyle factors and newly diagnosed diseases including colorectal carcinoma. The follow-up rate was more than 90% for each follow-up questionnaire cycle in the three cohort studies. Archival formalin-fixed paraffin-embedded (FFPE) tissue blocks of tumor and normal colon were collected in a subset of CRCs, using the prospective cohort incident-tumor biobank method^13^. WES on tumor-normal paired DNA from FFPE tissues was performed as described^14^ Clinical and somatic mutation data for 835 CRCs were downloaded from the Data Coordination Center (DCC) data portal at https://dcc.icgc.org/releases/current/Projects/COCA-CN; https://dcc.icgc.org/releases/current/Projects/COAD-US and https://dcc.icgc.org/releases/current/Projects/READ-US (as of January 2022). For consistency, only whole-exome sequencing (WES) samples (*n* = 295 samples in COCA-CN and *n* = 540 in COAD-US and READ-US) were analyzed.

### Dietary data

As previously described^15^, dietary intake assessment in NHS/HPFS was carried out with food frequency questionnaires (FFQ) that were initially collected in 1980 for NHS and in 1986 for HPFS. The NHS relied on a 61-item semiquantitative FFQ used at baseline^16^ and later expanded to approximately 130 food and beverage items in 1984, 1986, and every 4 years thereafter. The HPFS cohorts’ baseline dietary intake ascertainment relied on a 131-item FFQ that was also used for updates generally every 4 years subsequently^17^. The validity of this method in assessing dietary intake was robustly established by using diet and plasma nutrient records^18,19,20^. In this study, the dietary data were based on the most recent pre-diagnosis reported intake for each patient.

Specifically, unprocessed red meat consumption was evaluated based on the intake of “beef or lamb as main dish,” “pork as main dish,” “hamburger,” and “beef, pork, or lamb as a sandwich or mixed dish.”. Processed meat diets included “bacon”; “beef or pork hot dogs”; “salami, bologna, or other processed meat sandwiches”; and “other processed red meats such as sausage, kielbasa, etc.”. Participants reported their usual fruit and vegetable intake of standard portion size (e.g., 0.5 cups of strawberries, 1 banana, and 0.5 cups of cooked spinach) of fruit and vegetables for the preceding year of each FFQ. Frequencies and portions of fruit and vegetable items were converted to the average daily intake for each participant as previously described^21^. Consumption of red meat, poultry, fish, fruits and vegetables was evaluated in grams per day. Smoking status (never smoking, past smoking, current-smoking) and alcohol consumption (women: 0–<0.15, 0.15–<2.0, 2.0–<7.5, and ≥7.5 g/day; men: 0 to <1, 1–<6, 6–<15, and ≥15 g/day) were both stratified into three categories as previously described^5^.

### Germline data and *rs16906252*-T detection in the general population and patients with CRC

Germline data for the general population were retrieved (as of January 2022) from the 1000 Genomes Project Phase 3: http://grch37.ensembl.org/Homo_sapiens/Variation/Population?db=core;r=10:131265045-131266045;v=rs16906252;vdb=variation;vf=48812926; and the Genome Aggregation Database (gnomAD) v.2.1.1: https://gnomad.broadinstitute.org/variant/10-131265545-C-T?dataset=gnomad_r2_1 The *rs16906252*-T variant is in a promoter region. Promoter regions can be subject to low coverage in WES data. We thus discarded tumor/normal pairs with less than four reads covering the *rs16906252*-T locus in both samples, which roughly corresponds to a 95% statistical power to detect *rs16906252*-T if present (probability to detect for a heterozygous *rs16906252*-T patient is (1 − 0.5^4^). This filter resulted in *n* = 826 passing samples for NHS/HPFS (8% discarded samples) and *n* = 378 passing samples in (30% discarded samples). The higher number of discarded pairs in TCGA was mainly attributed to the use of older WES capture kits, such as SureSelect Human All Exon 38 Mb v2.

For the remaining samples, we used DeepVariant (version 1.4.0)^22^ to detect patients harboring *rs16906252*-T. This variant was considered germline if detected in tumor but not covered in the matched normal. There were no cases where both tumor and normal were covered with the tumor harboring *rs16906252*-T and not the matched normal sample. We also rescued *rs16906252*-T carriers by manually reviewing this locus in the Integrated Genomics Viewer^23^.

### Quantitative DNA methylation and microsatellite status analysis

MGMT promoter methylation analysis in the NHS/HPFS cohorts was carried out using bisulfite conversion and real-time PCR as previously described^24^. Microsatellite Instability (MSI) status was evaluated using 10 microsatellite markers (D2S123, D5S346, D17S250, BAT25, BAT26, BAT40, D18S55, D18S56, D18S67, and D18S487) as formerly detailed^12^.

### Average signature weights and cancer effect weights

The average signature weight (**Figure 2A**) is the average contribution of each mutational process to the total mutational burden in each tumor genome. Average cancer effect weights (**Figure 2B**) quantify the average contribution of each mutational process to the total cancer effect of variants classified as drivers in each tumor^25^. Cancer effects were quantified by estimation of underlying gene-based and site-based mutation rates and analysis compared to variant prevalence to quantify post-mutation cell proliferation and survival, as previously described^25^. To attribute cancer effects to signatures, effects of individual variants attributed to a signature were normalized to the total cancer effect of variants in each tumor. We then summed up the normalized cancer effects from all tumors and divided the sum by the total number of tumors.

### CRC ancestry determination

WES-derived ancestry calls from TCGA colorectal samples were downloaded from the National Cancer Institute Genomic Data Commons https://gdc.cancer.gov/about-data/publications/CCG-AIM-2020. The ancestry of each TCGA patient was determined using EthSeq^26^, which leverages genotype data from individuals of known ethnicity to infer the ancestry admixture of inputted individual genotypes. We applied this approach to NHS/HPFS samples to annotate their ethnicity (*n* = 900) based on DeepVariant germline variant calls from the normal samples. EthSeq-inferred results were highly concordant with self-determined ethnicities in NHS/HPFS: 4 out of 4 patients self-determined as Asian American were detected as such by EthSeq. In addition, 11 out of 13 patients who self-identified as African American were detected as such by EthSeq.

### GTEX data analysis

The data used for the analyses described in this manuscript were obtained at https://www.gtexportal.org/home/snp/rs16906252 from the GTEx Portal on 10/09/2022. In GTEx, the *P* values (**Figure S1A**) were generated by testing the alternative hypothesis that the slope of a linear regression model between genotype and expression deviates from zero. The normalized effect size (NES) of the eQTLs is defined as the slope of the linear regression and is computed as the effect of *rs16906252*-T relative to the reference allele.

### Mutational signature analysis

Mutations were classified by their 96 trinucleotide contexts for each sample. We then used the NMF R package and SigProfilerExtractor for the denovo signature extraction. While the NMF package performs a standard Kullback-Leibler-based non-negative matrix factorization directly on the mutational matrix^5^, SigProfilerExtractor resamples and normalizes it separately for each replicate^27^. Using their respective quality measures, rank 4 was selected as the best rank for both methods (Figure S8).

Of note, all the signatures mentioned in this manuscript were also found in WES. For the refitting, MutationalPatterns and SigProfilerAssignment were used. Both methods solve this non-negative least-squares (NNLS) optimization problem, using the pracma package and a custom implementation of the forward stagewise algorithm respectively^28,29^. Additionally, the decompose_fit function of SigProfilerAssignment was used to perform the decomposition of extracted signatures into their exome renormalized COSMIC components.

When compared to reference COSMIC v3 Single Base Substitution (SBS) signatures^30^, the four *de novo* signatures extracted using the NMF package displayed the highest similarity with SBS5 [cosine similarity (cossim) = 0.90], a featureless signature with unknown aetiology; SBS1 (cossim = 0.94), the age signature; SBS10a (cossim = 0.88) a POLE exonuclease deficiency signature; and SBS44 (cossim = 0.93) a defective DNA mismatch repair signature (**Figure S10**). Similarly, the 4 *de novo* signatures extracted using SigProfilerExtractor displayed the highest similarity with SBS5 (cossim = 0.888), SBS1 (cossim = 0.95), SBS10a (cossim = 0.873), and SBS44 (cossim = = 0.866) (data not shown).

### Genome-Wide Association Study (GWAS) data

The *rs16906252 MGMT* variant was found to associate with cancer risk in a GWAS study of DNA repair genes^7^ comprising 11,114 CRCs and 14,659 controls from the Colon Cancer Family Registry (CCFR) and the Genetics and Epidemiology of Colorectal Cancer Consortium (GECCO). Briefly, the authors leveraged 15,419 single nucleotide variants (SNVs) from 185 DNA repair genes and evaluated the association with CRC risk with a Binomial Sequential Goodness of Fit (BSGoF) procedure to adjust *P* values. Here, we replotted the SNPs from DNA repair genes as a Manhattan plot (**Figure S1B**) where we show the adjusted *P* values (*y*-axis) along the genome (position index, *x*-axis).

Fine-mapping of MGMT (top right inset, **Figure S1B)** was performed with LDassoc^31^. Both *P* values and *rs16906252*-T linkage disequilibrium patterns are shown for *MGMT* SNPs across all populations in the 1000 Genome Project.

### Statistical analysis, including regression models and interaction analyses

To investigate the interaction between *rs16906252*-T *MGMT* SNP and pre-diagnosis total (processed and unprocessed) red meat intake, we used a standard generalized linear model from R package ‘stats’ (version 4.2.1)^32^. In addition, we used a Tobit regression model^33^ as implemented in the R Package ‘AER’ (version 1.2-10)^34^. In the Tobit model, the observed range of the dependent variable is clustered at a lower boundary (**Figure S16**). Here, we used a left boundary of 0.01 and a right boundary of 0.99.

For the interaction analysis, we assume that *rs16906252*-T amplify the amount of DNA alkylating damage but does not directly cause it. Interaction contrasts were computed with the emmeans R package^35^.

We used R version 4.1.0 to perform statistical analyses. Significance for two-group comparisons was evaluated by one-sided Mann–Whitney U tests unless otherwise indicated. *P* < 0.05 was considered statistically significant.

## Results

### Prevalence of *rs16906252*-T in the general population and CRC

We first examined the prevalence of *rs16906252 C>T* (both CT and TT, hereafter simply referred to as *rs16906252*-T carriers) in the general population by leveraging data from the 1000 Genomes Project^36^ and the Genome Aggregation Database^37^ (gnomAD; **Figure S2**). We found that *rs16906252*-T was present in 12% of the study populations: 380 *rs16906252-*TT and 9174 *rs16906252-*CT versus 79,068 *rs16906252-*CC individuals in gnomAD. Strikingly, *rs16906252*-T was largely absent in individuals with African and East Asian ancestries and more abundant in individuals with European ancestry (**Figure S3**). When stratified by populations, *rs16906252*-T genotype proportions conformed to Hardy-Weinberg equilibrium (right-tail chi-square *P* > 0.05 after Bonferroni correction, **Table S1**).

We then detected (see **Methods**) and surveyed the distribution of *rs16906252*-T in two independent whole-exome sequencing (WES) CRC datasets from US-wide cohorts, namely NHS/HPFS^5^ (*n* = 826) and TCGA^38^ (*n* = 378). We used DeepVariant, a deep convolutional neural-network approach, to call genetic variants. Out of the 1440 CRC TCGA and NHS/HPFS patients analyzed, 132 were CT carriers and 20 were TT. The small sample size of TT individuals precluded their separate analysis, and we thus analyzed both CT and TT together (both CT and TT are hereafter simply referred to as rs16906252-T carriers). We found that overall, 12.4 % of patients with CRC harboured *rs16906252*-T (12.3% [= 102/826] of NHS / HPFS and 13.0% [= 49/378] of TCGA). *rs16906252*-T is significantly associated with *MGMT* promoter CpG island methylation in CRC^8^, including in the tumor specimens used in this analysis (Fisher’s exact tests *P* = 4.9 × 10^−8^ for TCGA, **Table S2,** *P* = 2.0 × 10^−13^ for NHS/HPFS, **Table S3** and *P* < 2.2 × 10^−16^ when aggregating TCGA and NHS/HPFS, **Figure 1A** and **Table S4**).

**Figure 1:**
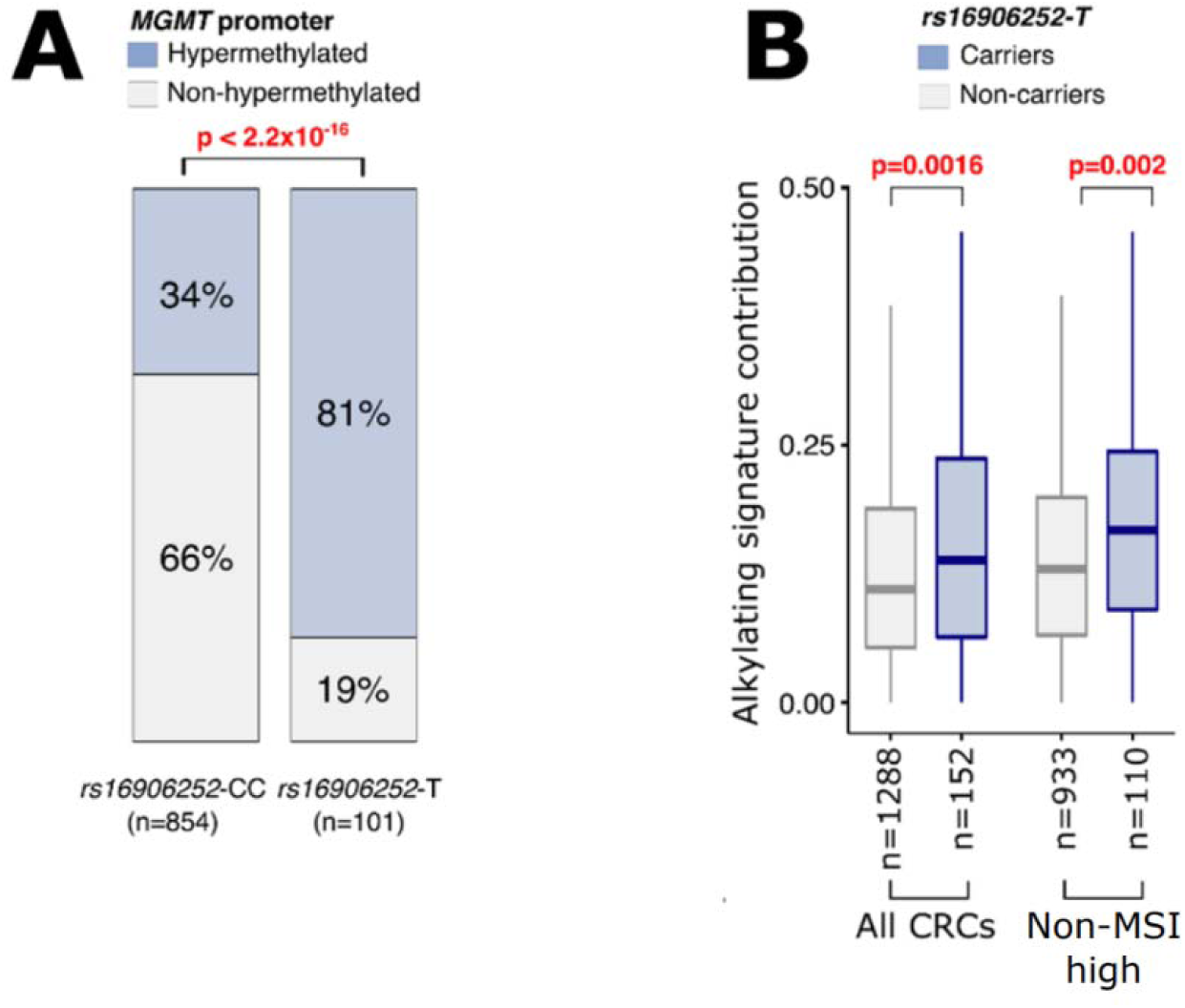
The *rs16906252*-T *MGMT* promoter variant is associated with alkylating damage in CRC. (**A**) Proportion of tumors with hypermethylated *MGMT* promoter stratified by *rs16906252*-T status. The Chi-square test *P* value is shown on top. (**B**) Proportion of mutations assigned to th alkylating signature in all CRCs and in non-MSI-high CRCs, segregated by *rs16906252*-T status in NHS/HPFS and TCGA. Mann-Whitney U test *P* values are shown. Box-plot outliers are not shown. CRC: Colorectal Cancer. Non-MSI-high: Non-microsatellite-high. NHS/HPFS: NHS, Nurses’ Health Studies I and II. HPFS, Health Professionals Follow-up Study. TCGA, The Cancer Genome Atlas.

**Figure 2:**
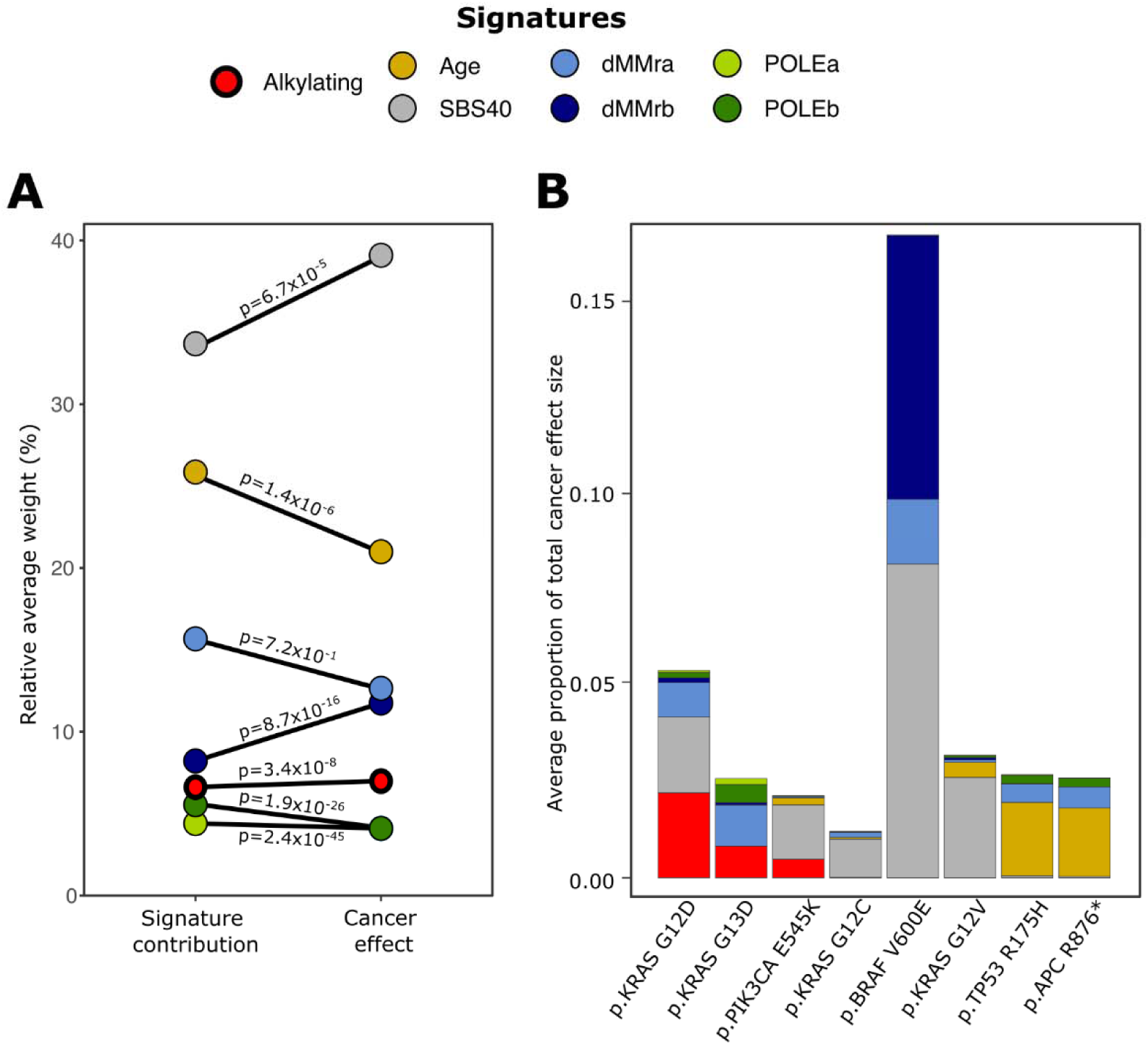
Average signature contribution and cancer effect weight in NHS/HPFS CRCs. (**A**) For each of the seven mutational signatures found in NHS/HPFS, the average contribution to the mutational burden (on the left) and the average contribution to the total cancer effect (on the right) is shown. Paired Wilcoxon Rank test *P* values for the difference of signature weight between TMB and cancer effect are also plotted. (**B**) The contribution of each signature to th average cancer effect of CRC hotspots. Here, the cancer effect is quantified proportionate to th total cancer effect of all recurrent variants in each tumor. dMMRa and dMMRb are the two signatures for DNA mismatch-repair deficiency. POLEa and POLEb are the two signatures for polymerase-epsilon deficiency. SBS40 is a featureless, flat signature with unknown aetiology.

### Tumors harboring *rs16906252*-T are enriched with alkylating damage

Because *MGMT* is a crucial gene for alkylating DNA damage repair^6^, we next investigated whether patients harboring *rs16906252*-T developed tumors that were enriched with the alkylating mutational signature^5^. TCGA and NHS/HPFS mutational signature deconvolution was performed as previously published by using a standard Non-Negative Matrix Factorization (NMF) approach^5,39^. After aggregating NHS/HPFS and TCGA, we found that CRC patients harboring *rs16906252*-T exhibit significantly higher levels of tumor alkylating damage among all CRCs and in non-microsatellite-high (non-MSI-high) CRCs (*P* = 1.6 × 10^−3^ and *P* = 2.0 × 10^−3^ respectively, Mann–Whitney U test, **Figure 1B**). In addition, we evaluated the effect size for the Mann–Whitney U tests by calculating the rank-biserial correlation *r_rb_*. We observed similar effect sizes for all CRCs and non-MSI-high CRCs only (*r_rb_* = 0.15 and 0.17 respectively). Tumors of NHS/HPFS CRC patients that were *rs16906252*-T carriers were enriched with alkylating damage (*r_rb_* = 0.21 and *P* = 3.2 × 10^−4^ among all CRCs; *r_rb_* = 0.21 and *P* = 9.2 × 10^−4^ in non-MSI-high CRCs, Mann–Whitney U test, **Figure S4A**). Tumors of TCGA CRC patients that were *rs16906252*-T carriers, as determined by DeepVariant, did not exhibit statistically significantly higher levels of alkylating damage (**Figure S4B**). However, after rescuing false-negative tumor/normal pairs with the Integrated Genomics Viewer (IGV, see **Methods** and **Figure S4C, Figure S4D,** and **Figure S4E**) by manual review, we observed increased alkylating damage among carriers in TCGA CRCs also (*P* = 0.075; *r_rb_* = 0.10 among all CRCs; and *P* = 0.043; *r_rb_* = 0.15 in non-MSI-high CRCs, Mann–Whitney U test, **Figure S4D**). Interestingly, we also observed that among tumors with *MGMT* promoter hypermethylation, there is a significantly higher amount of alkylating damage in tumors from rs16906252-T carriers (**Figure S5**). When we examined the association of the *MGMT* SNP with the other CRC mutational signatures [Age, mismatch repair deficiency (dMMR), POLE], we found no significant associations (**Figure S6**).

### Alkylating damage confers significant cancer advantage

The contribution of the mutational signatures to the total mutational burden is not necessarily equivalent to their contribution to oncogenic mutations^25^, but critically depends on the degree to which the subsequent mutations provide a selective advantage by increasing cancer cell proliferation or survival. Thus, we performed a direct calculation of cancer effect size as previously established^25^ that accounts for underlying gene– and site-based mutation rates to quantify both the selective advantage of each variant arising from the CRC mutagenic processes, as well as attribute these oncogenic effects to each mutational signature. This calculation quantifies the extent to which a mutational process accounts for carcinogenesis in a particular tumor. We observed that the alkylating signature contributes an average of 7% of the total oncogenic effect among non-MSI-high patients (**Figure 2A**) and that *KRAS* c.35G>A (p.G12D), *KRAS* c.38G>A (p.G13D), and *PIK3CA* c.1633G>A (p.E545K) were the main oncogenic variants targeted by the CRC alkylating signature (**Figure 2B**), consistent with our prior findings^5^. Indeed, our analysis demonstrated that 42% of the total carcinogenic effect of alkylating damage was mediated by key oncogenic mutations in *KRAS* p.G12D and p.G13D. Thus, the increased alkylating damage burden in *rs16906252*-T carriers can confer a substantial selective advantage to colorectal cancer cell lineages.

### Alkylating damage and *rs16906252*-T across ancestries

Because of the wide range of *rs16906252-T* frequencies across ancestries (**Figure 3A** and **Figure S3**), we investigated the contribution of CRC alkylating damage across ethnicities in NHS/HPFS, TCGA as well as COCA-CN, an independent cohort of Chinese patients with CRC (**Figure 3B**).

**Figure 3:**
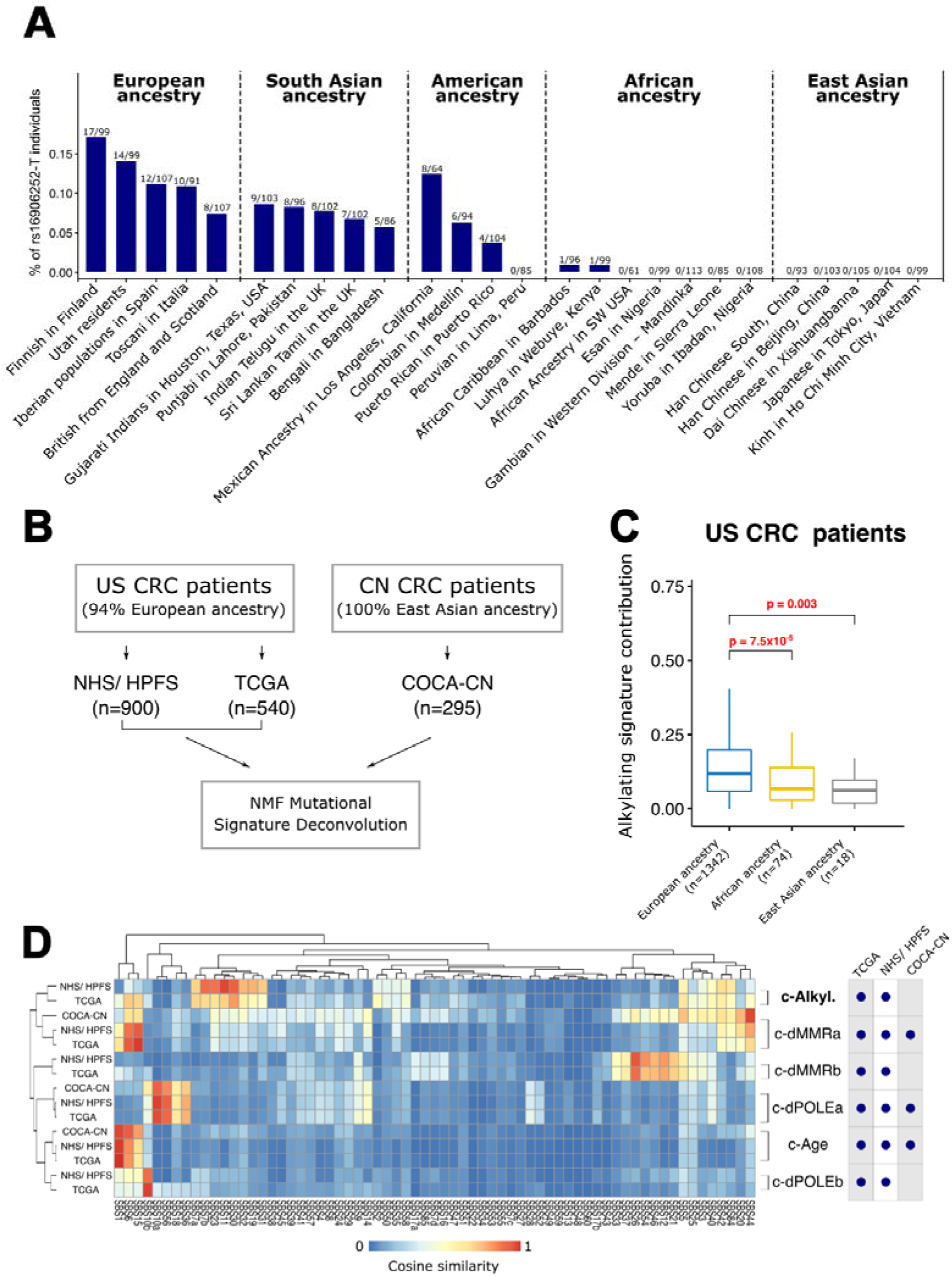
Tumor alkylating signature contribution across genetic ancestries. (**A**) Proportion of *rs16906252*-T carriers from the 1000 Genome project Phase 3. (**B**) Mutational signature deconvolution in US-wide cohorts (TCGA and NHS/HPFS, on the left) as well as one East Asian cohort (COCA-CN, on the right) (**C**) The proportion of mutations assigned to alkylating damage in CRC, stratified by genetic ancestries in TCGA and NHS/HPFS. (**D**) Heat map of the similarity scores between colorectal tumor (from TCGA, NHS/HPFS and COCA-CN) signatures, clustered on the *y*-axis, and reference COSMIC signatures, clustered on the *x*-axis. Clustering has been performed according to cosine similarity. The table on the right summarizes the presence / absence of each mutagenic process (*y* axis) in each dataset (*x* axis).

First, we investigated previously published WES-derived genetic ancestries in TCGA^40^ (*n* = 13 patients with East Asian ancestry and *n* = 42 patients with African ancestry). We similarly inferred genetic ancestries in NHS/HPFS using EthSeq^26^ (*n* = 5 patients with East Asian ancestry and *n* = 13 patients with African ancestry, see **Methods**). Overall, we observed a significant depletion in the alkylating signature in patients with East Asian (*r_rb_*= 0.26, *P* = 3.0 × 10^−3^, Mann–Whitney U test) and African ancestries (*r_rb_*= 0.38, *P* = 7.5 × 10^−5^, Mann–Whitney U test) compared to patients with European ancestry (**Figure 3C**). Comparable results were observed in TCGA (**Figure S7A**), but not in NHS/HPFS (**Figure S7B**) likely because of the relative lack of patients with non-European ancestry in the latter (14% and 2% patients with non-European ancestry in TCGA and NHS/HPFS respectively).

Next, we performed a mutational-signature analysis on COCA-CN (*n* = 295 samples, see **Methods** and **Figure 3D** and **Figure S8, Figure S9, Figure S10**). In contrast to TCGA and NHS/HPFS CRCs, the alkylating signature was absent in COCA-CN. We assessed the robustness of this result with three different methods. First, we extracted seven de novo signatures in COCA-CN and showed that none matched the alkylating signature (see **Methods** and **Figure S11**). Next, to demonstrate that the difference in sample size between COCA-CN (*n* = 295) and NHS/HPFS (*n* = 900) cannot explain the lack of alkylating signature detection in COCA-CN, we (i) randomly sampled 295 CRCs of the 900 from NHS/HPFS; (ii) extracted seven signatures from the 295 CRCs; and (iii) repeated steps (i) and (ii) a hundred times. In 72% of the simulations, either SBS11 or SBS30^5^ was detected (**Figure S12**); we previously found these two signatures to be interchangeable in smaller sample-size cohorts because of their similarity (cossim = 0.81) ^5^. Lastly, we fitted COCA-CN mutational profiles to SBS signatures found in TCGA-CRCs (SBS1, SBS10a, SBS10b, SBS30, SBS15, SBS26 and SBS40) using the MutationalPatterns R package^29^. The age (SBS1) and noise signatures (SBS40) showed no significant difference between TCGA and COCA-CN. The dMMR (SBS15 and SBS26) and POLE (SBS10a and SBS10b) signature contributions were significantly lower in COCA-CN compared to TCGA, due to the lower proportion of MSI / POLE patients in COCA-CN (0.7% POLE deficient and 3.8% MSI samples as computed by MSIseq^41^ compared to TCGA (1.8% POLE deficient and 14.0% MSI samples). The alkylating signature (SBS30) was significantly lower in COCA-CN (*P* < 2.2 × 10^−16^) compared to TCGA CRCs (**Figure S13**). We further demonstrated the lack of detectable signature in COCA-CN by decomposing COCA-CN signatures into combination sets of COSMIC reference signatures and showing the lack of an alkylating signature contribution (**Figure S14 A**). In addition, no alkylating signature was found in COCA-CN after deconvoluting signatures for other ranks (rank 2 to 15 shown on **Figure S14 B**). Altogether, these results support the lack of a detectable alkylating mutational signature in COCA-CN, consistent with the lower frequency of *rs16906252*-T in individuals of East Asian ancestry.

### *rs16906252*-T and red meat intake potentiate alkylating damage

The addition of nitrates (e.g. during processing) and catalysis by heme iron in red meat can lead to the formation of N-nitroso compounds (NOCs), which are alkylating agents^15,42,43^. We previously demonstrated an association of pre-diagnosis processed and unprocessed red-meat intake with the CRC alkylating mutational signature in NHS/HPFS^5^. This previous analysis relied on the comparison of extreme eaters (top 10%) to the rest of the cohort. Here, we leveraged a more general approach of regression modeling that did not rely on discretization of a continuous variable into binary data. To find lifestyle/ dietary behaviors associated with a change in alkylating damage, we also further expanded our analysis to include prospectively collected repeated measurements of smoking and alcohol, in addition to dietary variables of fruit, vegetable, chicken, and fish consumption in the NHS and HPFS cohorts (see **Methods**). Because the distribution of alkylating damage appears to be truncated normally distributed with zero-inflation (**Figure S15**), we used a Tobit regression model^44^ which showed that among all variables studied, only red meat is associated with CRC alkylating damage (*P* = 0.019, **Figure 4A**). We observed similar results with a standard Generalized Linear Model (GLM; *P* = 0.014, **Figure S16A**).

**Figure 4:**
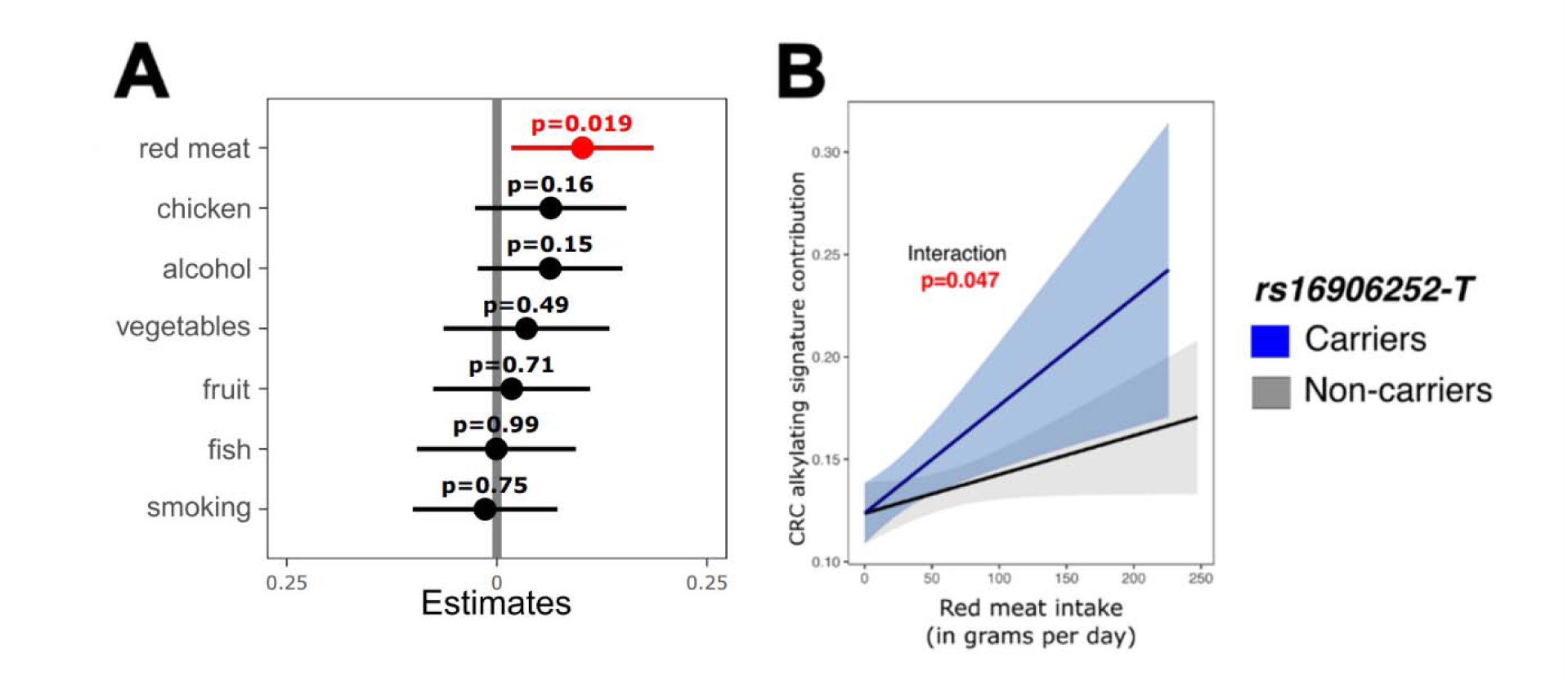
Effects of lifestyle and *rs16906252*-T variant for CRC alkylating damage in NHS/HPFS. (**A**) Tobit model of dietary variable and alkylating damage in CRC after *z*-scoring (centering and scaling of all the variables) (**B**) Interactive effect and 95% confidence intervals of *rs16906252*-T and pre-diagnosis red meat consumption on CRC alkylating signature contribution using a Tobit model (solid lines). NHS/HPFS: NHS, Nurses’ Health Studies I and II. HPFS, Health Professionals Follow-up Study.

Next, we tested for an interaction between *rs16906252*-T and pre-diagnosis red-meat intake in a prediction model of alkylating damage. We observed a significant interaction of red meat intake with the tumor alkylating mutational signature for *rs16906252*-T carriers (interaction-term Wald test *P* = 0.046 in the Tobit Model, **Figure 4B**). Similar results were observed in the GLM model (interaction-term Wald test *P* = 0.053, **Figure S16B**).

## Discussion

Our study demonstrates that the *rs16906252*-T *MGMT* promoter germline variant associates with a CRC alkylating signature that has a substantial carcinogenic effect, especially by originating cancer driver mutations *KRAS* p.G12D and p.G13D^5^. This result is consistent with prior studies showing a link between *MGMT* loss and *KRAS* c.35G>A^10,45^. In addition, we observe that among tumors with *MGMT* promoter hypermethylation, there is a significantly higher amount of alkylating damage in tumors from rs16906252-T carriers. We thus postulate that individuals with the rs16906252-T germline variant may have accumulated augmented alkylating damage over the course of their lifetime.

In addition, we demonstrated a negligible contribution of alkylating damage to tumor mutational burden and to mutagenic oncogenesis in an East Asian CRC cohort, which can be partly attributed to the paucity of *rs16906252*-T in this population. This is consistent with a previously described weaker association between red meat intake and CRC in East Asian populations^46^, however, large East Asian prospective or retrospective studies combined with genomic datasets would enable confirmation of the contribution of diet differences to the lack of a detectable alkylating signature. Alkylating damage arises from a wide array of modifiable exposures^47,48^ including red-meat intake, which in our study was the only variable significantly associated with alkylating damage among all the dietary variables studied. We observe a synergistic gene-by-environment interaction effect of the *rs16906252*-T germline variant and high pre-diagnosis red-meat intake, leading to increased tumor alkylating damage. Consistently, experimental studies have previously shown that *MGMT*-null mice develop tumors only in the presence of alkylating exposures^49,50^. Overall, our results support a causal relationship between red meat intake and CRC and encourage the implementation of precision prevention by enhanced screening and/or dietary modifications for individuals harboring the *rs16906252*-T variant.

## Acknowledgements

We thank G. Wang and S.McGrath for technical feedback, as well as S. Gazal and N. Oak for useful comments. The authors would like to acknowledge the contribution to this study from central cancer registries supported through the Centers for Disease Control and Prevention’s National Program of Cancer Registries (NPCR) and/or the National Cancer Institute’s Surveillance, Epidemiology, and End Results (SEER) Program. Central registries may also be supported by state agencies, universities, and cancer centers. Participating central cancer registries include the following: Alabama, Alaska, Arizona, Arkansas, California, Delaware, Colorado, Connecticut, Florida, Georgia, Hawaii, Idaho, Indiana, Iowa, Kentucky, Louisiana, Maine, Maryland, Massachusetts, Michigan, Mississippi, Montana, Nebraska, Nevada, New Hampshire, New Jersey, New Mexico, New York, North Carolina, North Dakota, Ohio, Oklahoma, Oregon, Pennsylvania, Puerto Rico, Rhode Island, Seattle SEER Registry, South Carolina, Tennessee, Texas, Utah, Virginia, West Virginia, Wyoming.

This work was supported by Cancer Research UK Grand Challenge Award (UK C10674/A27140 to M.G. and S.O.); by the U.S. National Institutes of Health (NIH) grants R01 CA151993 (to S.O.) and R35 CA197735 (to S.O.); by the American Cancer Society Clinical Research Professor Award (Grant # CRP-24-1185864-01-PROF to S.O.); and by the Project P and Crush Colon Cancer Funds. M.G. was supported by a High Pointe Investigatorship in Gastrointestinal Oncology. The Nurses’ Health Study was supported by the NIH grants UM1 CA186107 and P01 CA879690. The Nurses’ Health Study II was supported by the NIH grant U01 CA176726. The Health Professionals Follow-up Study was supported by NIH grants P01 CA055075, UM1 CA167552, and U01 CA167552.

## Data Availability Statement

Publicly available data used in this study was accessed from the Data Coordination Center data portal at <u>COCA-CN</u>, <u>COAD-US</u>, and <u>READ-US</u>; from the 1000 Genomes Project Phase 3 at <u>rs16906252</u>; from the Genome Aggregation Database (gnomAD) v.2.1.1 at <u>10-131265545-C-T</u>; from the National Cancer Institute Genomic Data Commons at <u>CCG-AIM-2020</u>; and from the GTEx Portal at <u>rs16906252</u>. NHS/HPFS WES data have been deposited in dbGAP at accession number <u>phs000722</u>. All analysis scripts are available upon request.

## Author Contributions

CG: Conceptualization, Data curation, Formal Analysis, Investigation, Methodology, Validation, Visualization, Writing – original draft, Writing – review & editing; JC: Formal Analysis, Writing – review & editing; CT: Formal Analysis, Writing – review & editing; BR: Methodology, Writing – review & editing; MH: Methodology, Writing – review & editing; TU: Data curation, Writing – review & editing; JAM: Resources, Writing – review & editing; JAN: Writing – review & editing; ELG: Resources, Writing – review & editing; JPT: Methodology, Writing – review & editing; SO: Funding Acquisition, Resources, Methodology, Validation, Writing – review & editing; MG: Conceptualization, Data curation, Funding acquisition, Investigation, Methodology, Project administration, Resources, Supervision, Writing – original draft, Writing – review & editing.

