## Supplemental Table 1 for "Germline predisposition to oncogenic alkylating damage in colorectal cancer"

**Table S1: Hardy-Weinberg equilibrium analyses for rs16906252-T across gnomAD populations**

|  | **C/C** | **C/T** | **T/T** | ***P* value** | **Bonferroni adjusted *P* value** |
| --- | --- | --- | --- | --- | --- |
| **European (Finnish)** | 7861 | 1483 | 75 | 0.85903 | 1 |
| **European**  **(non-Finnish)** | 30845 | 5050 | 217 | 0.80434 | 1 |
| **Ashkenazi Jewish** | 3653 | 563 | 18 | 0.75777 | 1 |
| **Others** | 2252 | 319 | 18 | 0.19988 | 1 |
| **South Asian** | 10157 | 862 | 32 | 0.01157 | 0.10413 |
| **Latino/Admixed American** | 11304 | 749 | 18 | 0.31723 | 1 |
| **African/African American** | 7397 | 147 | 2 | 0.35129 | 1 |
| **East Asian** | 5599 | 1 | 0 | 0.9999 | 1 |
