## Supplemental Table 2 for "Germline predisposition to oncogenic alkylating damage in colorectal cancer"

**Table S2: The *rs16906252-T* *MGMT* promoter variant and tumor *MGMT* promoter hypermethylation in TCGA CRCs**

Fisher’s exact test *P* = 4.9 × 10^−8^

|  | No *rs16906252*-T | *rs16906252*-T |
| --- | --- | --- |
| Unmethylated MGMT promoter | **307** | **11** |
| Methylated MGMT promoter | **164** | **36** |
