## Supplemental Table 3 for "Germline predisposition to oncogenic alkylating damage in colorectal cancer"

**Table S3: The *rs16906252-T* *MGMT* promoter variant and tumor *MGMT* promoter hypermethylation in NHS / HPFS**

Fisher's exact test *P* = 2.0 × 10^−13^

|  | No *rs16906252*-T | *rs16906252*-T |
| --- | --- | --- |
| Unmethylated MGMT promoter | **260** | **8** |
| Methylated MGMT promoter | **123** | **46** |
