## Supplemental Table 4 for "Germline predisposition to oncogenic alkylating damage in colorectal cancer"

**Table S4: The *rs16906252-T* MGMT promoter variant and tumor *MGMT* promoter hypermethylation in NHS/ HPFS and TCGA CRCs**

Fisher's exact test *P* < 2.2 × 10^−16^

|  | No *rs16906252*-T | *rs16906252*-T |
| --- | --- | --- |
| Unmethylated MGMT promoter | **567** | **19** |
| Methylated MGMT promoter | **287** | **82** |
