## Supplemental Figure 1 for "Germline predisposition to oncogenic alkylating damage in colorectal cancer"

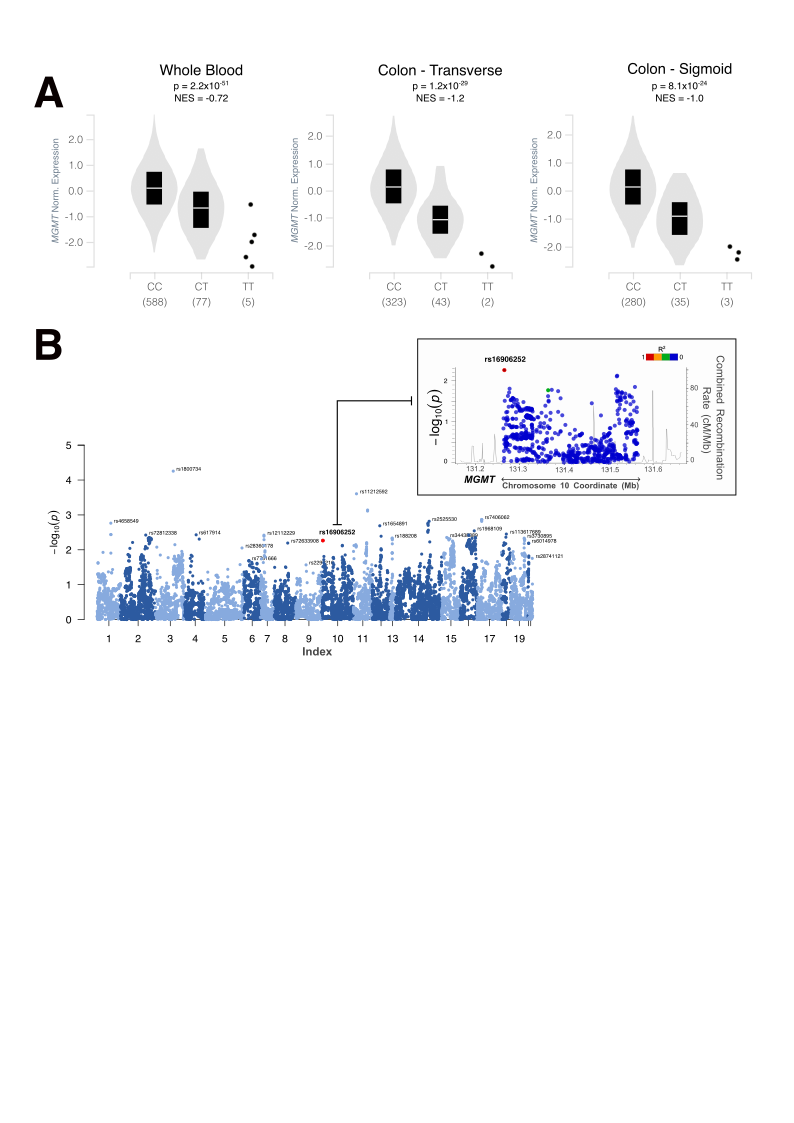

**Figure S1: The rs16906252-T MGMT promoter variant is a cis-eQTL**

(**A**) The rs16906252-T *MGMT* promoter variant and *MGMT* gene expression levels from the Genotype-Tissue Expression (GTEx) project in Blood (left panel), Transverse colon (middle panel) and Sigmoid colon (right panel). Rectal tissue data was not available in GTEx. *P* values and Normalized Effect Size (NES) measures were retrieved from the GTEx portal and are detailed in **Methods.** (**B**) Association of CRC risk with SNPs in DNA repair genes in the Colon Cancer Family Registry (CCFR) and Genetics and Epidemiology of Colorectal Cancer Consortium (GECCO) studies. The inset (top right) shows the fine mapping of *MGMT* SNPs.
