## Supplemental Figure 2 for "Germline predisposition to oncogenic alkylating damage in colorectal cancer"

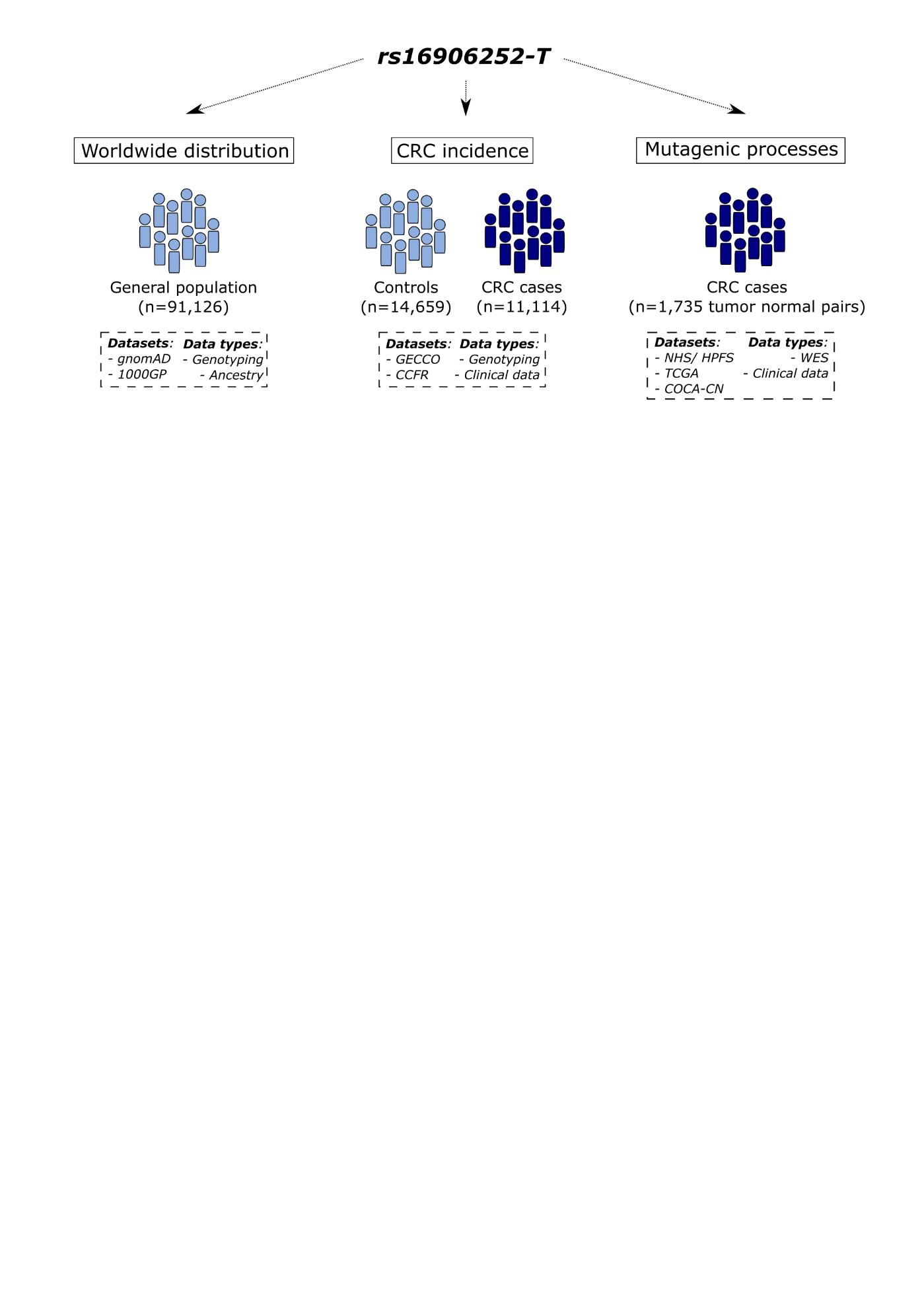

**Figure S2: Data overview**

Datasets used: 1000GP, 1000 Genomes Project. gnomAD, Genome Aggregation Database. NHS, Nurses' Health Studies I and II. HPFS, Health Professionals Follow-up Study. TCGA, The Cancer Genome Atlas. COCA-CN, Colorectal adenocarcinoma in China. CCFR, Colon Cancer Family Registry. GECCO, Genetics and Epidemiology of Colorectal Cancer Consortium.
