## Supplemental Figure 3 for "Germline predisposition to oncogenic alkylating damage in colorectal cancer"

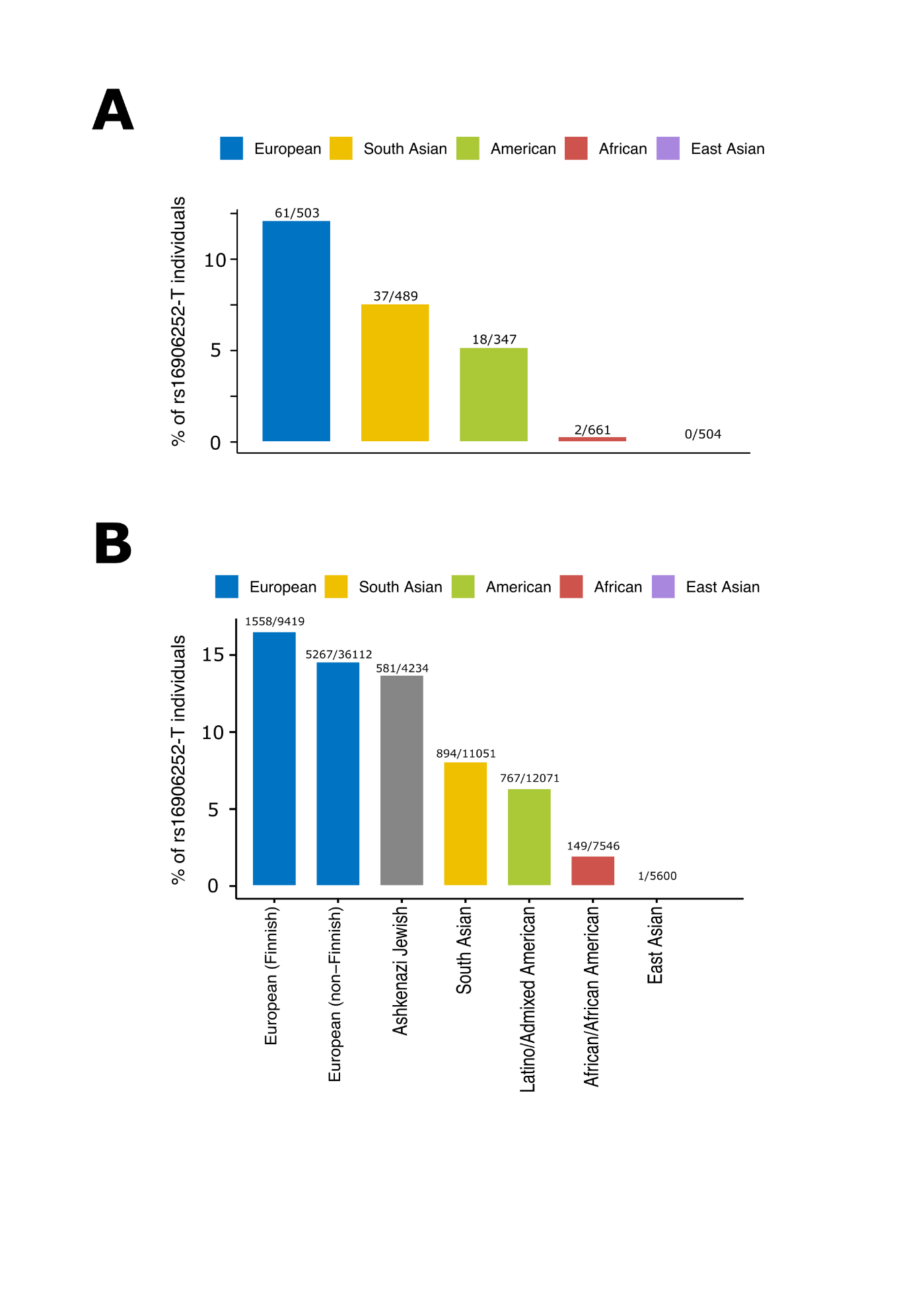

**Figure S3: Distribution of *rs16906252*-T across populations.**

Proportion of *rs16906252*-T individuals (both homozygous and heterozygous) from (**A**) The 1000 Genome Project Phase 3 and (**B**) The Genome Aggregation Database (gnomAD).
