## Supplemental Figure 4 for "Germline predisposition to oncogenic alkylating damage in colorectal cancer"

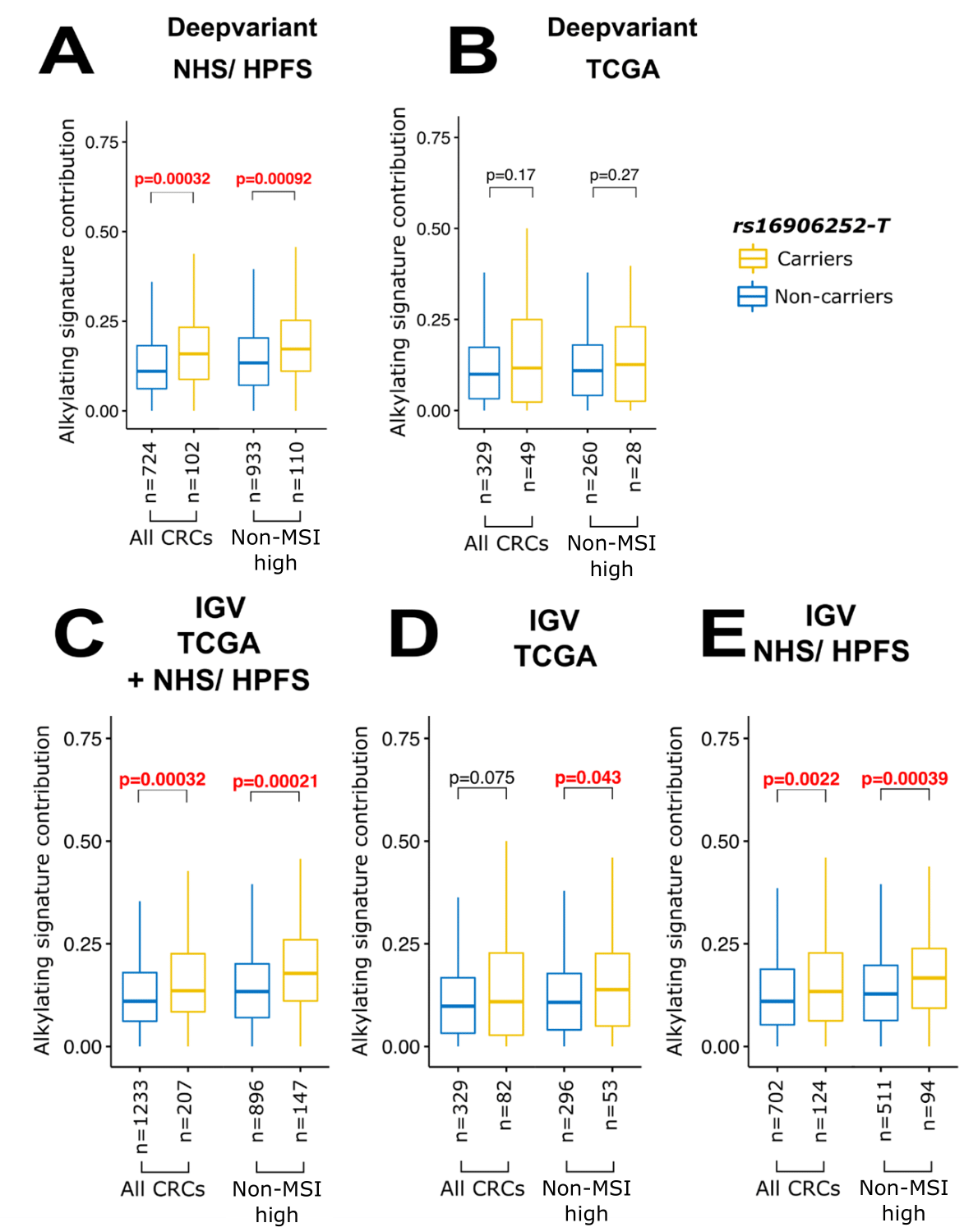

**Figure S4: The *rs16906252*-T *MGMT* promoter variant and CRC alkylating damage**

Proportion of mutations assigned to the alkylating signature in CRC segregated by *rs16906252*-T status as detected by DeepVariant in (**A**) NHS/ HPFS CRCs (**B**) TCGA CRCs. We also show the same analysis but with *rs16906252*-T status detected with IGV manual curation in (**C**) Both NHS/HPFS and TCGA CRCs (**D**) TCGA CRCs only (**E**) NHS/ HPFS CRCs only. Box-plot outliers are not shown. NHS, Nurses' Health Studies I and II. HPFS, Health Professionals Follow-up Study. TCGA, The Cancer Genome Atlas. IGV: Integrative Genomic Viewer.
