## Supplemental Figure 5 for "Germline predisposition to oncogenic alkylating damage in colorectal cancer"

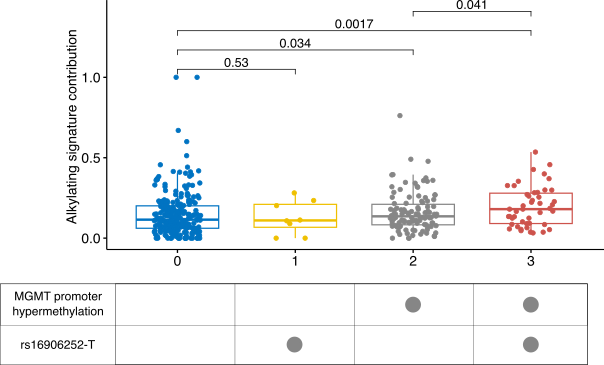

**Figure S5: CRC alkylating damage stratified by rs16906252-T and *MGMT* promoter hypermethylation status**

Proportion of mutations assigned to the alkylating signature in CRC (y axis) segregated by *rs16906252*-T status and *MGMT* promoter hypermethylation (x axis). The table on the bottom summarizes the presence/ absence of rs16906252-T/ *MGMT* promoter hypermethylation.
