## Supplemental Figure 6 for "Germline predisposition to oncogenic alkylating damage in colorectal cancer"

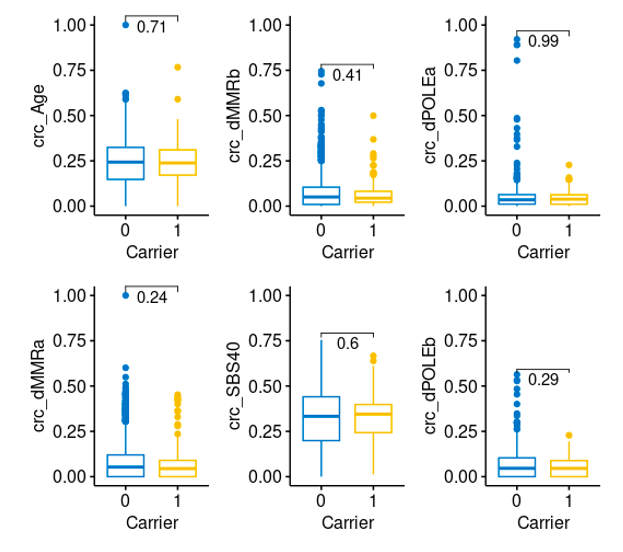

**Figure S6: Association of CRC mutational signatures with rs16906252-T status**

Mutational signature contribution (y axis) in non-carriers and carriers of rs16906252-T (x axis, 0 and 1 respectively).
