## Supplemental Figure 7 for "Germline predisposition to oncogenic alkylating damage in colorectal cancer"

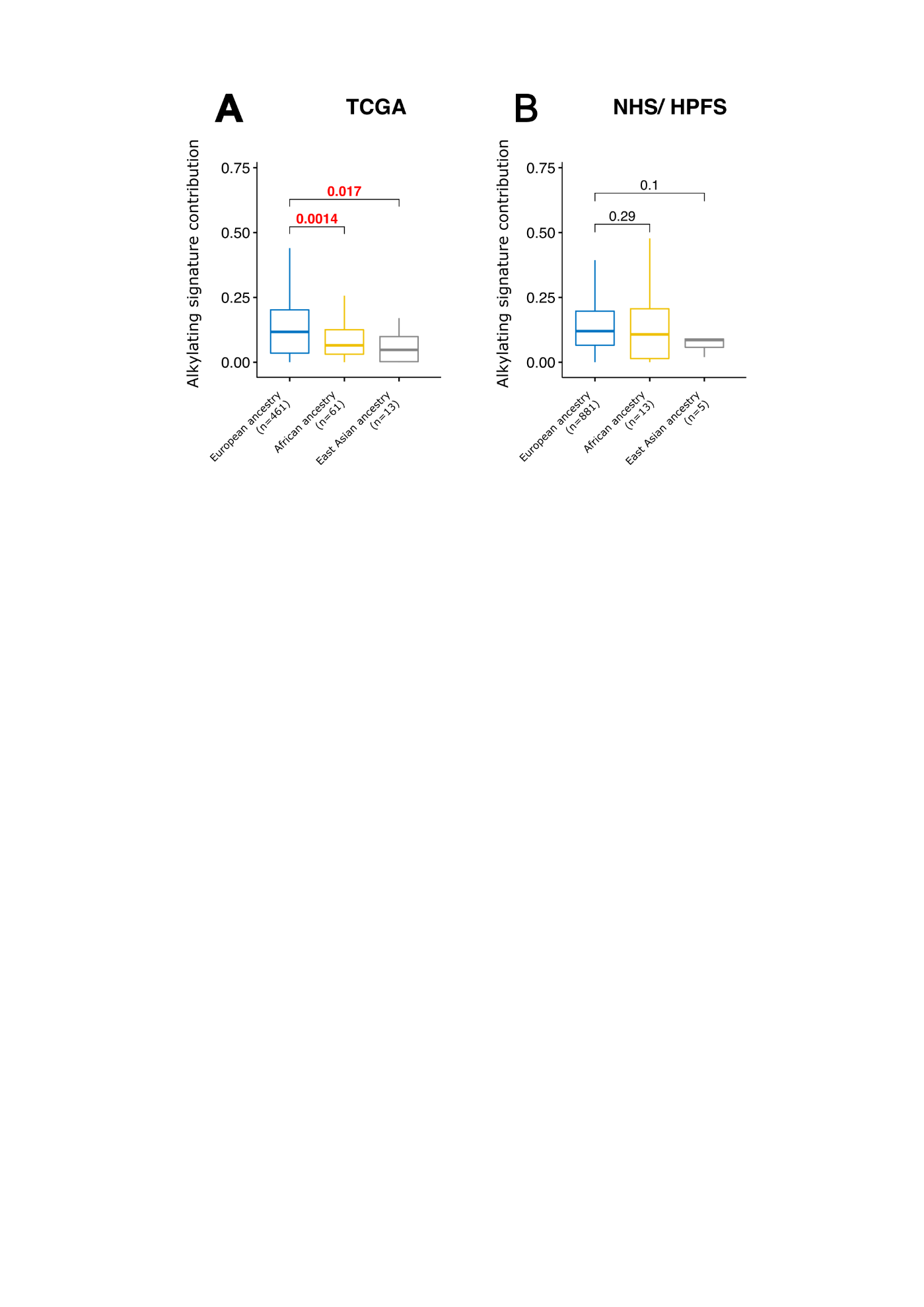

**Figure S7: Alkylating signature contributions to CRC across genetic ancestries in TCGA and NHS/ HPFS patients.**

The proportion of mutations assigned to alkylating damage in CRC, stratified by genetic ancestries in (**A**) TCGA and (**B**) NHS/ HPFS. Box-plot outliers are not shown. NHS, Nurses' Health Studies I and II. HPFS, Health Professionals Follow-up Study. TCGA, The Cancer Genome Atlas.
