## Supplemental Figure 8 for "Germline predisposition to oncogenic alkylating damage in colorectal cancer"

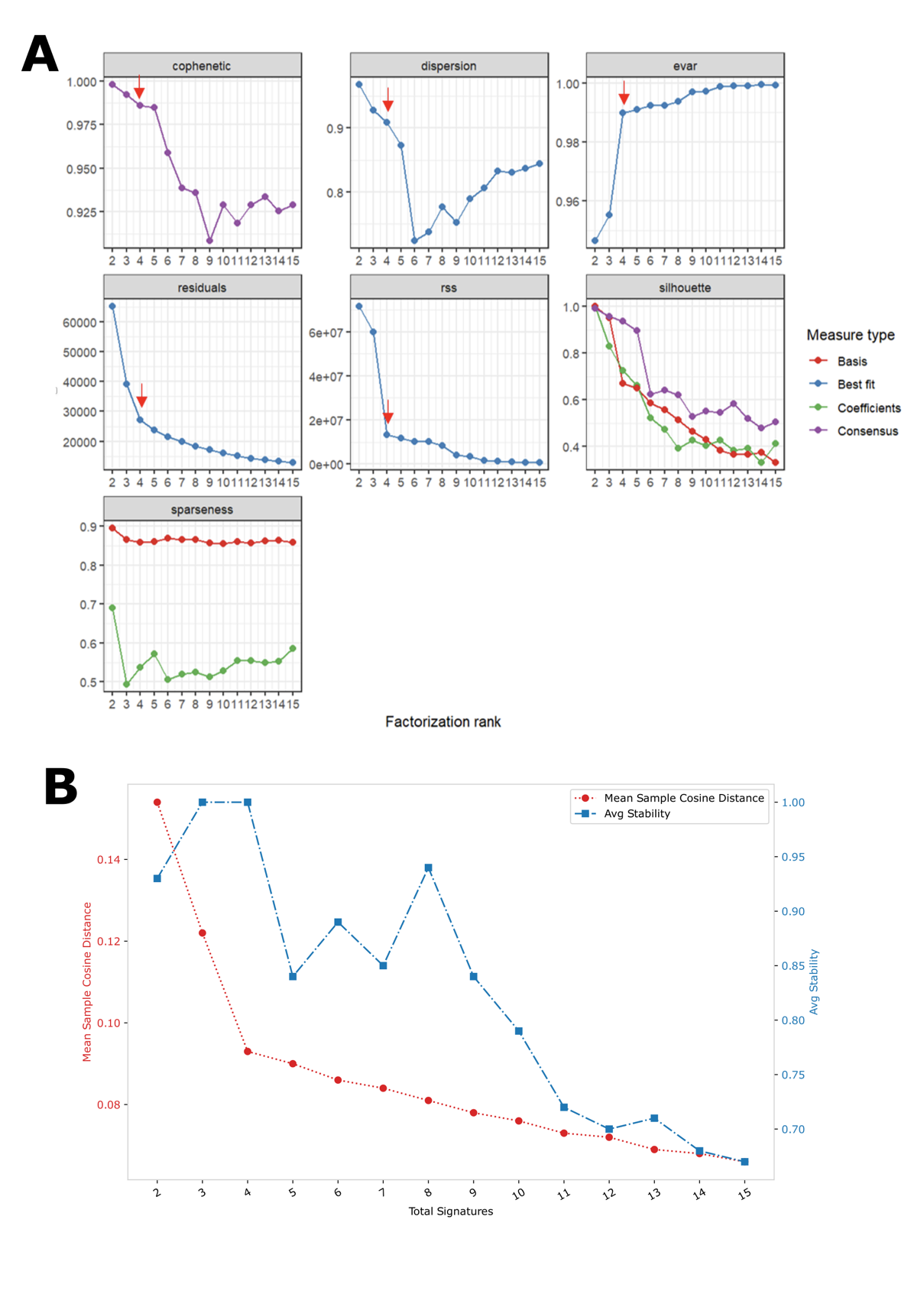

**Figure S8: Non-negative matrix factorization rank survey in COCA-CN CRCs.**

Quality measures for non-negative matrix factorization in COCA-CN, as described in the (**A**) R package "NMF" and (**B**) SigProfilerExtractor tool. Arrows indicate the estimated rank of mutational signatures.
