## Supplemental Figure 9 for "Germline predisposition to oncogenic alkylating damage in colorectal cancer"

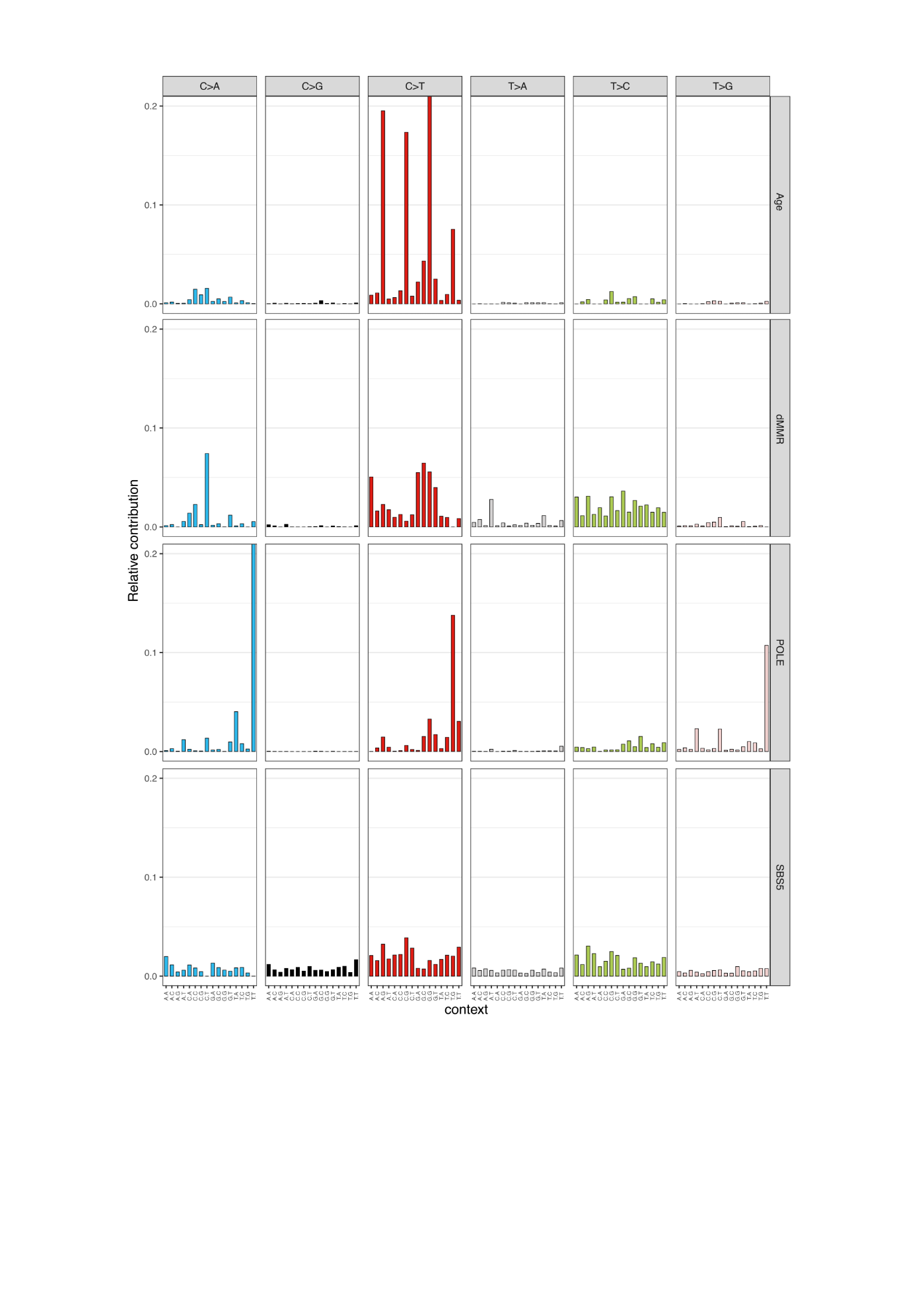

**Figure S9: De novo signature deconvolution in COCA-CN CRCs.**

The consensus four signatures found by NMF in COCA-CN.
