## Supplemental Figure 10 for "Germline predisposition to oncogenic alkylating damage in colorectal cancer"

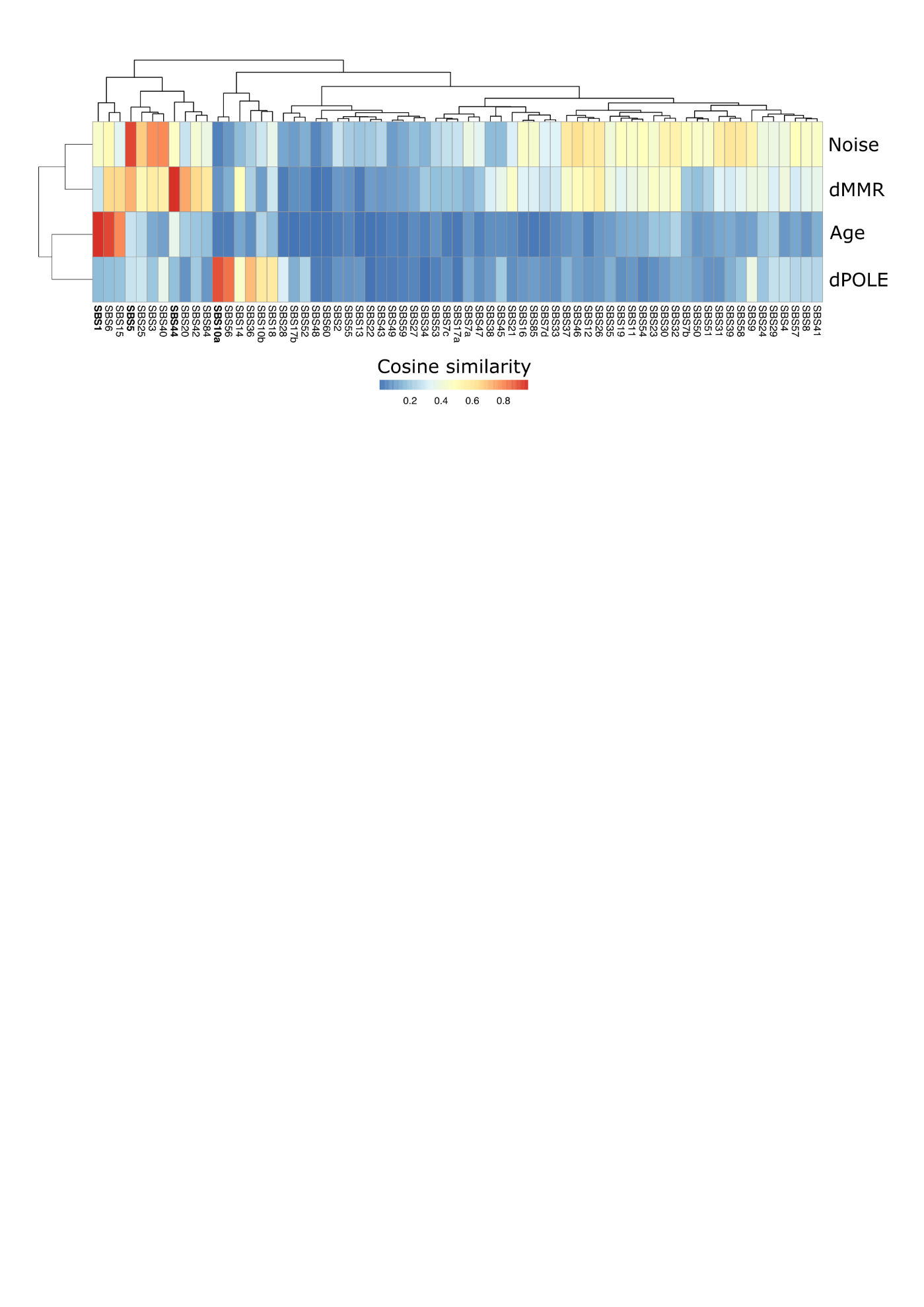

**Figure S10: Comparison between four COCA-CN CRC signatures and COSMIC signatures.**

Cosine similarity heat map between four de novo signatures found in COCA-CN (*y* axis) and COSMIC v3 signatures (*x* axis). COCA-CN signatures were named after their closest COSMIC mutational process match (bold). COCA-CN: Colorectal adenocarcinoma in China.
