## Supplemental Figure 11 for "Germline predisposition to oncogenic alkylating damage in colorectal cancer"

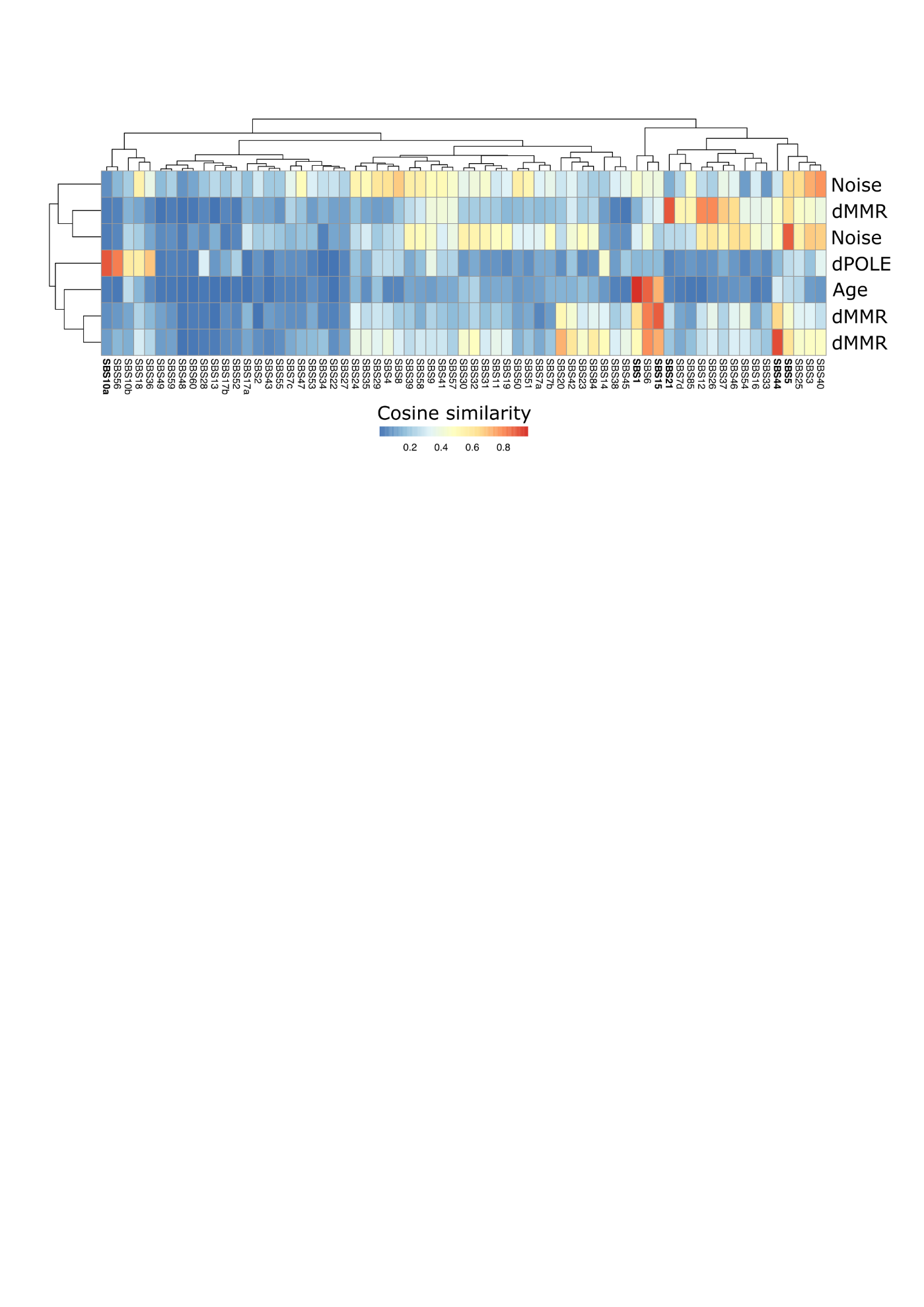

**Figure S11: Comparison between seven COCA-CN CRC and COSMIC signatures.**

Cosine similarity heat map between seven de novo signatures found in COCA-CN (*y* axis) and COSMIC v3 signatures (*x* axis). COCA-CN signatures were named after their closest COSMIC v3 mutational process match (bold). COCA-CN: Colorectal adenocarcinoma in China.
