## Supplemental Figure 12 for "Germline predisposition to oncogenic alkylating damage in colorectal cancer"

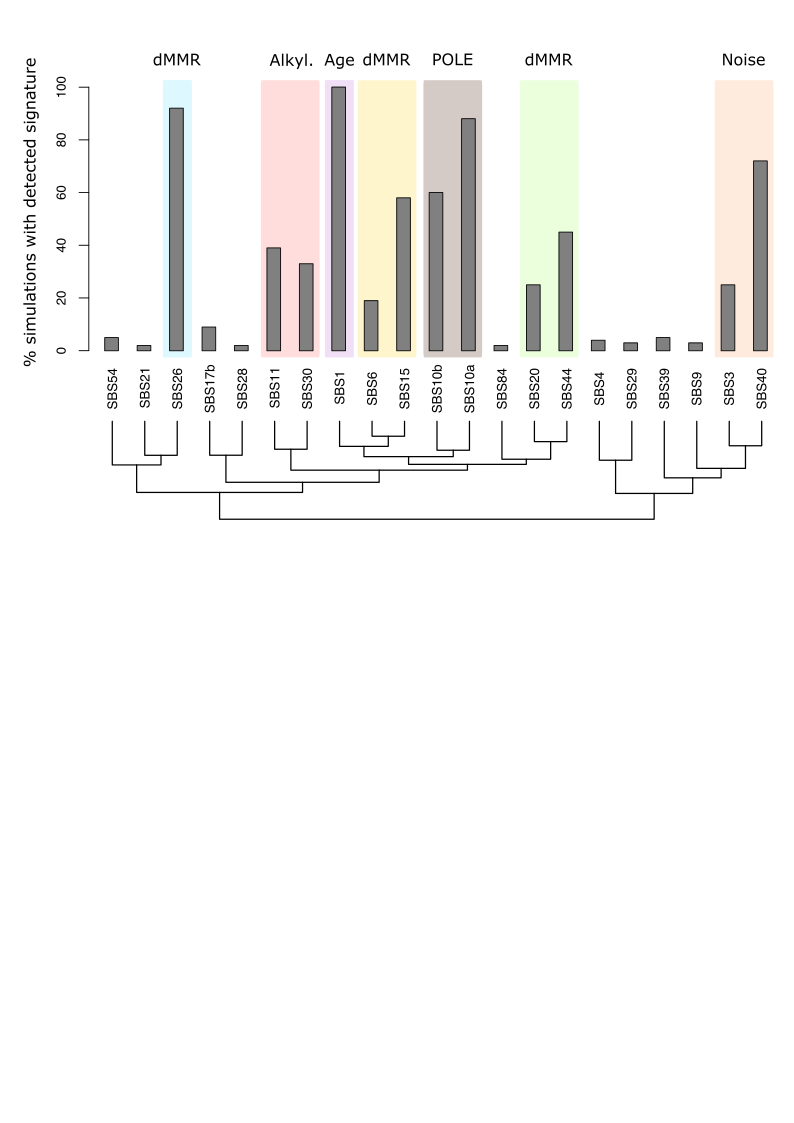

**Figure S12: Signature assignment after under-sampling simulations from 900 (NHS/HPFS sample size) to 295 (COCA-CN CRC sample size).**

The proportion of each unique mutational signature deconvoluted from 100 under-sampling simulations is shown. After under-sampling to 295 samples, signatures can be attributed to the ones found in the original 900 NHS/ HPFS tumors (SBS1, SBS11, SBS26, SBS10a, SBS10b, SBS15 and SBS40) or to a signature with similar aetiology/cosine similarity (colored boxes). Hierarchical clustering of signatures based on their cosine similarity is shown on the *x*-axis.
