## Supplemental Figure 13 for "Germline predisposition to oncogenic alkylating damage in colorectal cancer"

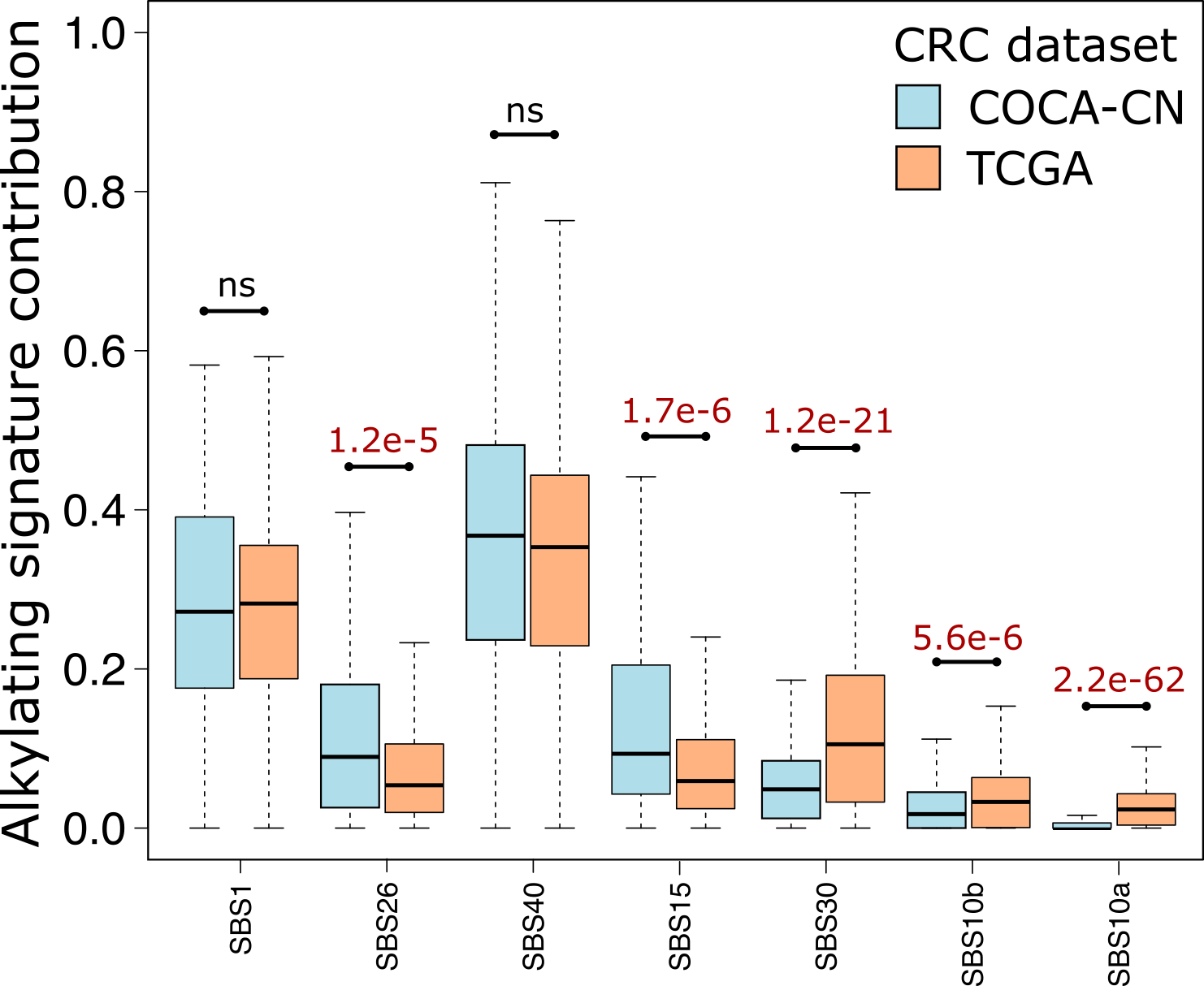

**Figure S13: Signature-fitting approach for TCGA and COCA-CN.**

De novo signature extraction in TCGA and signature fitting in COCA-CN CRCs using SBS signatures identified in TCGA (SBS1, SBS10a, SBS10b, SBS15, SBS26, SBS30 and SBS40). COCA-CN: Colorectal adenocarcinoma in China. TCGA, The Cancer Genome Atlas.
