## Supplemental Figure 14 for "Germline predisposition to oncogenic alkylating damage in colorectal cancer"

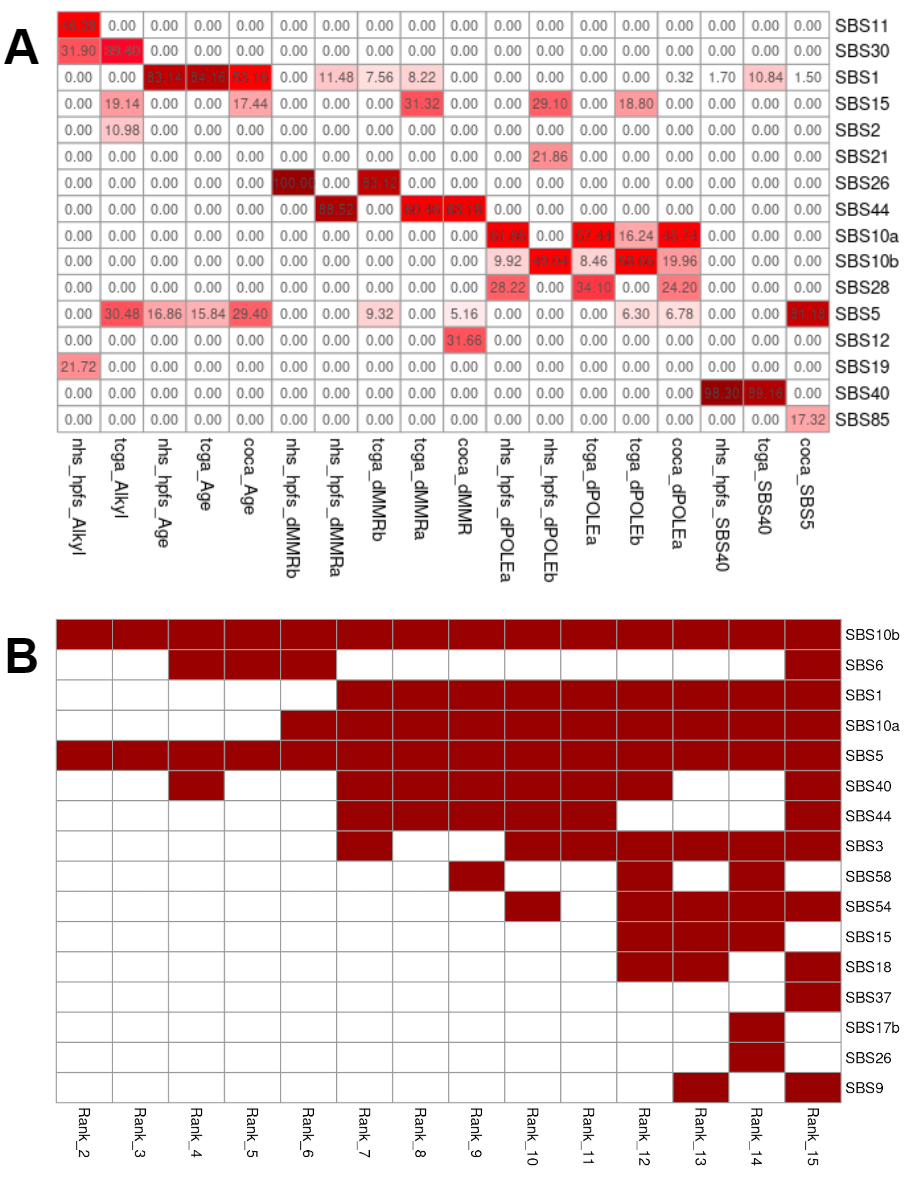

**Figure S14: Signature decomposition for all extracted signatures and additional ranks for COCA-CN extracted signatures**

(A) Decomposition of de novo extracted mutational signatures from NHS/HPFS, TCGA, and COCA-CN cohorts (x axis) to their COSMIC mutational signatures components (y axis), using SigProfilerAssignment’s decompose_fit function. De novo extracted signatures names are composed of the cohort they originated from and their closest COSMIC v3 mutational process match. (B) Map of de novo extracted mutational signatures from ranks 2-15 in COCA-CN cohort using NMF to their closest COSMIC mutational signature match (using cosine similarity).
