## Supplemental Figure 15 for "Germline predisposition to oncogenic alkylating damage in colorectal cancer"

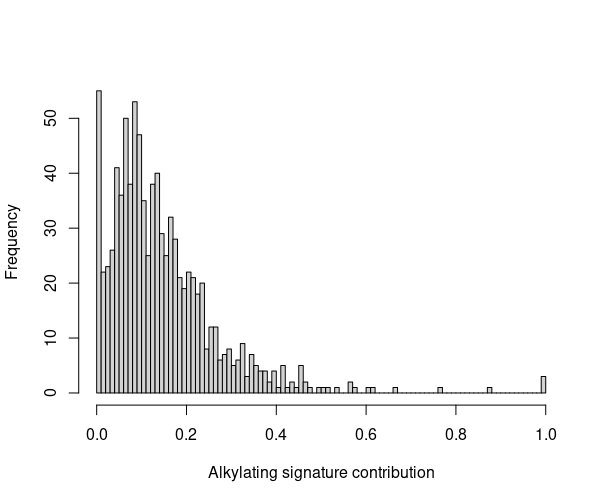

**Figure S15: Histogram of tumor alkylating damage in NHS/ HPFS.**

Distribution of alkylating signature contribution in NHS/ HPFS CRCs (*n* = 900). NHS / HPFS: NHS, Nurses' Health Studies I and II. HPFS, Health Professionals Follow-up Study.
