## Supplemental Figure 16 for "Germline predisposition to oncogenic alkylating damage in colorectal cancer"

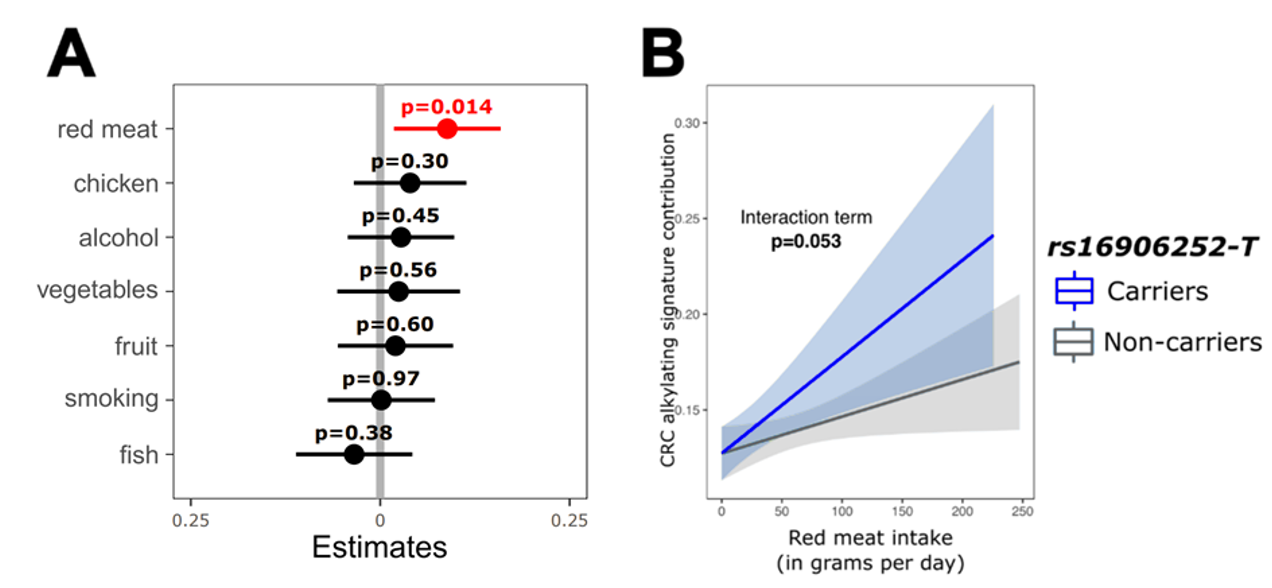

**Figure S16: Effects of lifestyle and *rs16906252*-T variant for CRC alkylating damage in NHS/ HPFS**

(**A**) Generalized linear regression model of dietary variables and alkylating damage in CRC after z-scoring (centering and scaling of all the variables) (**B**) Interactive effect and 95% confidence intervals of *rs16906252*-T and pre-diagnosis red meat consumption on CRC alkylating signature contribution using a Generalized linear regression model (solid lines). NHS / HPFS: NHS, Nurses' Health Studies I and II. HPFS, Health Professionals Follow-up Study.
